# The *global TB portfolio model*: a tool for projecting the epidemiological impact of TB policy options

**DOI:** 10.1101/2025.05.17.25327819

**Authors:** Sandip Mandal, Srinath Satyanarayana, Finn McQuaid, Peter J. Dodd, Nicolas A. Menzies, Richard G. White, Nimalan Arinaminpathy, Rein M. G. J. Houben, David W. Dowdy, Mikaela Smit, Suvanand Sahu, Carel Pretorius

## Abstract

**Background:** Tuberculosis (TB) remains one of the deadliest infectious diseases globally. Despite the World Health Organization’s (WHO) End TB Strategy targets for 2035, progress has been hindered by structural, financial, and implementation barriers, including recent cuts in global funding. Strategic use of mathematical modelling is useful for prioritizing high-impact interventions and optimizing limited resources. A new global TB infection transmission model was developed to address limitations in existing tools with respect to these applications.

**Methods:** The model includes enhanced features such as age-specific mixing, explicit representation of asymptomatic TB, stratification by drug resistance, HIV status, and new vaccine status, and inclusion of both public and private care pathways. It was calibrated to country-specific data using Bayesian adaptive Markov Chain Monte Carlo (MCMC) methods. The model was used to assess the impact of national strategic plans and the Global Plan to End TB, using a Target Population (TP) component to map interventions to WHO guidelines.

**Results:** Model calibration showed good agreement with historical TB data from 29 high-burden countries, with case studies for Indonesia and Nigeria presented here. In Indonesia, comprehensive implementation of Global Plan interventions - including public-private mix efforts, modern diagnostics, improved treatment for drug-resistant TB, and a post-exposure vaccine - could enable the country to achieve End TB targets by 2035. In Nigeria, implementing its National Strategic Plan could reduce TB incidence by 27% and mortality by 37% by 2030, even without a vaccine. The model highlighted the additional efforts that are needed to meet the End-TB goals.

**Conclusions:** The enhanced TB model provides a flexible, policy-relevant framework for assessing the epidemiological impact of TB interventions at both national and global levels. Its open-source design and alignment with WHO recommendations make it a valuable tool for guiding evidence-based investments amid tightening global health budgets.

## Introduction

Tuberculosis (TB) remains one of the deadliest infectious diseases globally, posing a significant public health challenge despite concerted efforts to control and eliminate it. In 2014, the World Health Organization (WHO) introduced the End TB Strategy, setting ambitious targets to reduce TB incidence and mortality by 90% and 95%, respectively, by 2035, compared to 2015 levels.^1^ However, achieving these targets has proven to be exceedingly difficult due to various structural, financial, and implementation barriers. A critical challenge in this regard is the substantial funding gap that impedes the effective rollout of TB prevention, diagnosis, and treatment interventions.^2,3^ The recent reduction in US government funding for global TB programs has exacerbated this financial shortfall, further complicating efforts to scale up interventions in high-burden regions.^4–7^ In such a constrained resource environment, it is imperative to prioritize interventions that maximize impact. Additionally, a clear understanding of the magnitude of the funding required to meet specific TB control targets is essential for policymakers and global health organizations.

Mathematical modelling serves as a powerful tool in this context, enabling researchers and decision-makers to assess the impact of various TB interventions and allocate resources effectively.^8–12^ By simulating different scenarios, mathematical models can help predict the potential reduction in TB incidence and mortality associated with specific interventions, such as intensified case detection, improved treatment adherence, or vaccine deployment. These models also provide valuable insights into the cost-effectiveness of different strategies, allowing policymakers to optimize limited financial resources to achieve the maximum public health benefit. In light of existing financial constraints, leveraging mathematical modelling to guide evidence-based decision-making is crucial to closing the funding gap and advancing toward the global End TB targets.

TB infection transmission models have provided critical inputs to TB strategy development, at global and national level alike. The Stop TB Partnership (STB)’s Global Plans to End TB 2016-2020 and subsequent Global Plans for the periods 2018-2022, 2023-2030 and Global Fund’s Investment Case analysis towards the 5th to 7th replenishment,^13,14^ for example, were informed by results obtained from the TIME Impact model.^15^ However, while TIME Impact was widely used in many settings due to a comprehensive design it has the limitation that WHO guidelines and recommendations and STB’s Global Plan strategies cannot be clearly mapped to TIME Impact’s intervention structures. Its extensive use in modelling applications at country and global despite this key limitation (and other limitations detailed below), prompted calls from technical guidance groups to improve and redesign key aspects of the model to inform the Global Fund’s Investment Case for the 8^th^ replenishment, as the first in a planned series of key applications of the new model.

This paper outlines the methodology behind these improvements, presents validation results, and discusses the broader implications for TB control strategies and its facilitation through the newly introduced ‘Target Population’ component.

## Methodology

### Broadening global modelling impact methodology

The TIME Impact model is a user-friendly epidemiological transmission model designed to enhance local capacity building and support country-specific policy discussions.^15^ By providing national and subnational TB programs with a structured framework to analyse intervention impact, TIME Impact has played a critical role in informing funding applications—both at the governmental level (e.g., Ministry of Finance) and among international donors (e.g., the Global Fund to Fight AIDS, Tuberculosis and Malaria). Several case studies, for example its application to TB epidemics in South Africa and Ghana, demonstrated the utility of the TIME model for informing strategic planning and policy making by national TB programs.^15^

The TIME Impact model is nested within TIME, a broader suite of TB modelling tools integrated into Spectrum software, a widely used global health modelling platform available for free download^16^. The model’s flexibility and accessibility have enabled its application in diverse settings, where it has contributed to strategic decision-making and resource mobilization.

Despite its strengths, the current version of TIME Impact has limitations. One key limitation is its significant run time due to its detailed model structure and its integration into the Spectrum suite. Its runtime, which can be a few minutes per scenario (depending on runtime configuration settings) makes auto-fitting and optimization methods based on TIME Impact nearly impossible to implement, outside of cloud- and grid-computing environments. While not prohibitive in terms of country-level use – one of its main purposes – its runtime limitations proved too limiting for application to global level analysis requiring frequent multiple runs and many countries. Without a statistical inference framework, the model also lacks plausibility bounds for its results.

Methodological constraints include an assumption of homogeneous mixing, a common but simplified approach in TB modelling that does not fully account for heterogeneities in contact patterns, particularly age-specific mixing. The natural history structure lacks more recent developments, such as the inclusion of explicit health sates to account for asymptomatic TB. Its intervention structure lacks structure for non-NTP service provisions, which is critical in many high burden countries where the private healthcare sector plays a substantial role in managing TB. It does not include structure for large scale vaccination.

### A new TB transmission model

The Global Model Advisory Group was convened by the TB Modelling and Analysis Consortium (TB MAC), bringing together representatives from a range of organizations: including modelling experts affiliated with TB MAC, WHO, the Stop TB Partnership (STB), the Global Plan’s working group, the Global Fund’s guidance group, and others. The group met to discuss the limitations of existing tools for global-level TB analysis and to assess proposed improvements.

A new dynamical TB model was developed under the guidance of the advisory group to address these limitations and to meet emerging demands from the global modelling community (including those issued by STB’s and Global Fund’s guidance groups). It expands on the TIME model methodology in many respects, addresses its limitations, while maintaining good design agreement with models generally used for similar strategic modelling purposes.

Figure 1 presents a schematic representation of the model, which delineates the natural history of tuberculosis (TB) and incorporates key aspects of diagnosis, treatment, and the role of both public (NTP) and private (non-NTP) healthcare sectors in disease transmission. In 2024, WHO updated the definitions of symptomatic and asymptomatic TB to improve to enhance surveillance, diagnosis, and treatment strategies. Symptomatic TB includes individuals with typical TB symptoms who often seek care, while asymptomatic (or subclinical) TB refers to those without symptoms but who test positive for TB, either with or without bacteriological confirmation. Accordingly, we assumed, following infection, individuals may develop asymptomatic or symptomatic TB. Asymptomatic individuals are assumed to be less infectious than those with symptoms.^17,18^ Upon the onset of symptomatic TB, individuals typically seek care from either the public or private sector. Successful diagnosis leads to entry into the treatment cascade within the respective sector. However, if diagnosis fails or treatment is not initiated, individuals transition to missed diagnosis or temporarily disengaged states. These individuals may re-enter care pathways unless they self-cure or succumb to the disease.

**Figure 1.**
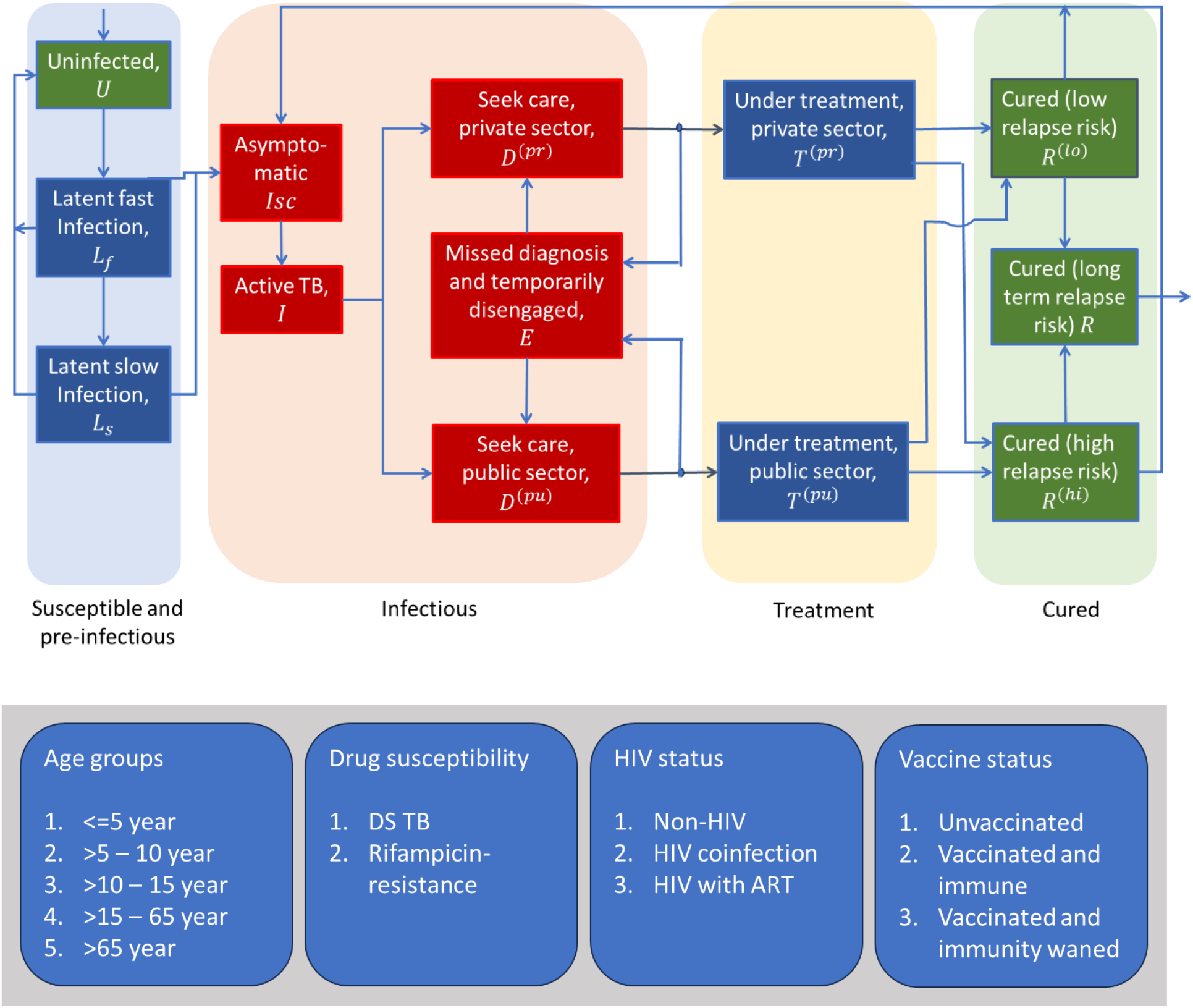
Schematic illustration of the model structure. Infectious compartments contributing to the force-of-infection are shown in red. For clarity, the diagram omits certain rates incorporated in the model, including self-cure; exogenous reinfection; and background mortality. Details of parameters and model equations are given in the supplementary document.

Post-treatment outcomes vary depending on treatment completion. Those completing treatment are considered cured with a low relapse risk, while individuals who are lost to follow-up during treatment enter a high relapse-risk-state. Those failing treatment are considered infectious. Relapses predominantly occur within the first two years following recovery, after which the remaining individuals assume a stabilized lifetime relapse risk.^19–21^ The model also accounts for reinfection, with reduced susceptibility across compartments having infectious history. Notably, most TB-related deaths occur prior to treatment initiation - primarily within the pre-treatment compartments, particularly in low case detection settings, indicated by red shading in the diagram, excluding asymptomatic individuals.

The model is stratified by age, drug susceptibility, HIV status, and vaccination status (vaccinated, non-vaccinated and vaccinated with waning immunity). Age stratification includes five groups: ≤5 years, >5–10 years, >10–15 years, >15–65 years, and >65 years. It incorporates an explicit age-contact matrix, allowing for age-mixing and a more nuanced representation of age-dependant transmission dynamics. The epidemiology of both drug-sensitive and rifampicin-resistant TB strains is represented within the model. Although HIV dynamics are not explicitly modelled, the impact of HIV on TB outcomes is addressed by categorizing all compartments into three groups: HIV-negative individuals, HIV-positive individuals not on antiretroviral therapy (ART), and HIV-positive individuals receiving ART. Additionally, the model accounts for new TB vaccination status by dividing the population into three categories: individuals who have never received vaccine, individuals who are vaccinated and remain immune, and individuals whose vaccine-derived immunity has waned.

Though the full model consists of 585 compartments, it excludes gender differences in TB burden, the distinction between pulmonary and extrapulmonary TB, and different risk groups (e.g. Diabetics, other comorbidities), and assumes an average infectiousness across these forms.

#### The Target Population component

The intervention modelling approach detailed above achieves most impact model needs but it does so without a clear correspondence to WHO guidelines for screening and TB care or to the recommendations of the TB Global Plan with respect to TB interventions. We developed a Target Population (TP) component to interface between the epidemiological impact model (the subject of this paper) and the resource needs estimation tool of the global TB portfolio model (not discussed in any detail here) to establish explicit conformance to WHO’s evolving patient-centred guidelines and to approve alignment between cost and impact models.

The TP component consists of 20 population groups (10 representing patient-initiated population groups and 10 representing provider-initiated population groups at risk for TB disease) to accommodate the necessary variations in WHO guidelines and recommendations for screening, diagnostic and treatment.^22^ The TP model processes a care cascade for each patient population, following closely the approach of WHO’s IHT TB model.^23^The alignment of the TP model’s patient-centred structure and with WHO recommendations for detection, treatment and prevention allows for integration of the model into the TB component of WHO’s Integrated Health Tool (IHT) as an impact modelling tool, allowing detailed country-level applications to be developed – a use of the model for which a case study is not yet available.

From these population-specific TB cascade structures three impact pathways are approximated with hazard rates, by aggregating over all populations: rates of detection (H_d_ in Figure 2) and treatment, population-level risks for developing TB disease (adjusted for higher risk in population groups receiving TPT, H_i_ in Figure 2), and treatment success rates for new and recurrent cases (H_tsr_ in Figure 2).

**Figure 2.**
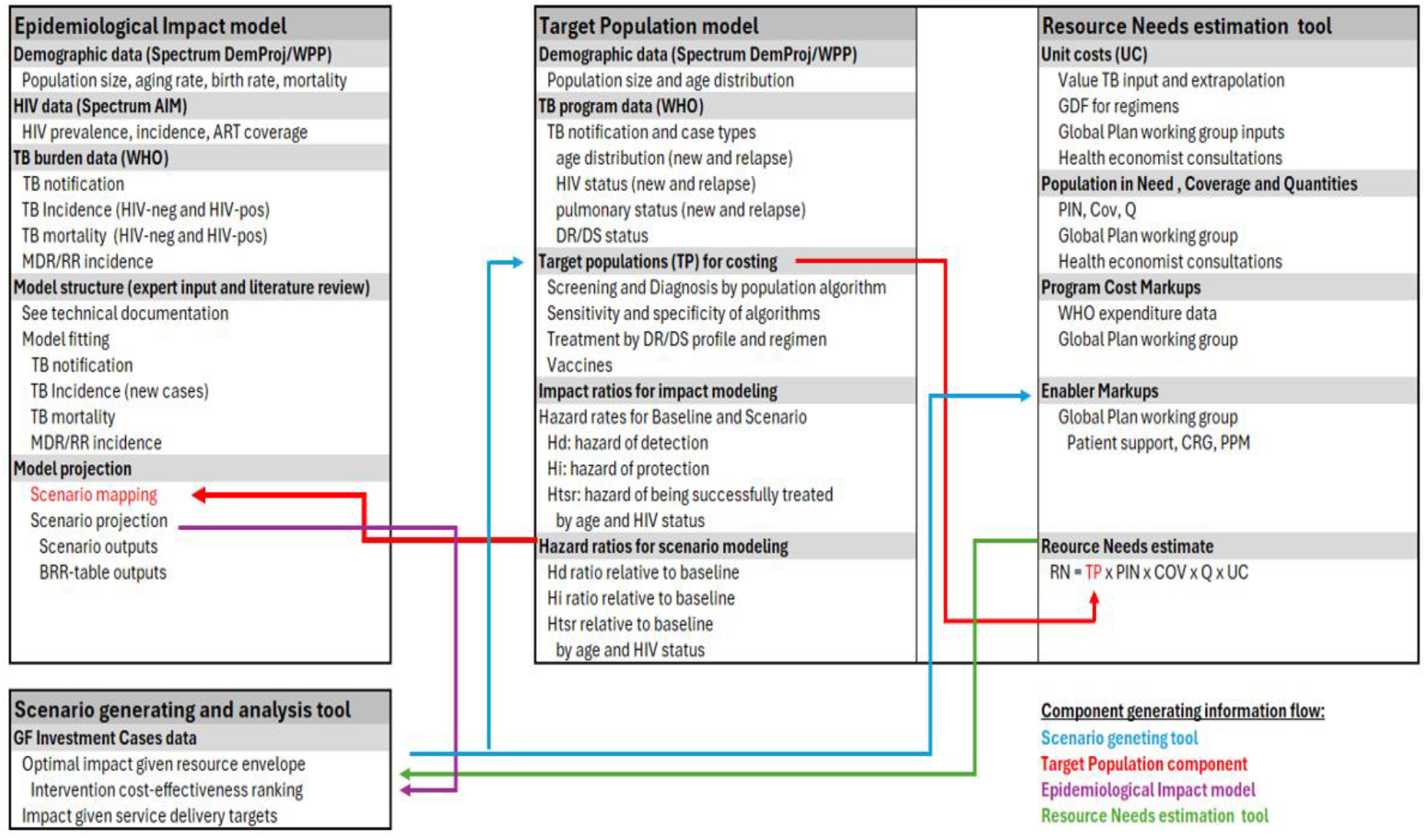
Schematic illustration of the flow of information between the Target Population model, the Epidemiological Impact model (the focus of this paper) and the Resource Needs estiamtion tool. RN-resource needs, PIN-population in need, COV-coverage, Q-quantitym UC-unit cost, BRR-benchmarking, reporting, and review.

To estimate the impact of the scenario projection, hazard ratios for a given scenario compared to a counterfactual, quantify increased detection, prevention and treatment outcomes are used to change impact related parameters in the model.

The flow of information between the Target Population model, the Epidemiological Impact model and the Resource Needs estimation tool is shown in Figure 2. In an application such Global Fund’s Investment Case analysis, a scenario-generating tool is used to populate the TP structure with intervention details (baseline and target coverage levels) corresponding to a specific scenario. Hazard ratios are applied to a calibrated epidemiological impact model and TP outputs (volumes or patient numbers requiring services needed for costing) are used by the Resource Needs estimation tool. Results from these models are used to produce optimal impacts of resource allocations, return on investment results and other outputs needed for a given analysis.

### Model calibration and credible intervals

The models were calibrated to 29 high-burden and Global Fund eligible countries (representing 90% of the TB burden in the Global Fund’s TB portfolio), with case studies presented here focussing on Indonesia and Nigeria, using the country specific data listed in Table 1. This was performed using Bayesian adaptive Markov chain Monte Carlo (MCMC), performing 50,000 iterations.^24^ We constructed the posterior density as follows: for each of the calibration targets described above, we constructed beta distributions to capture model proportions, and log-normal distributions to capture population rates, adjusting distribution parameters to match the central and uncertainty intervals of each calibration target. We also included wide prior uniform distributions for uncertain model parameters, such as the rates of treatment completion in the private sector. We then took the posterior density to be proportional to the product of all likelihood and prior densities. For practical purposes we calculated the log-posterior density, therefore taking a sum of the log-probability distributions of each of the individual likelihood components. After discarding the burn-in and taking a ‘thinned’ subsample, we drew 250 samples from the posterior density. Computing all model outputs (e.g. future incidence, mortality etc) using each of these samples, we estimated point values as the 50th percentile, and 95 percent Bayesian credible intervals (Crl) as being bounded by the 2.5th and 97.5th percentiles.

**Table 1:** Target data used for model calibration.

| Indicators |  | Sources |
| --- | --- | --- |
| All TB | Proportion of symptomatic TB among prevalent cases 2022 | General assumption- based on national TB prevalence survey in different countries<sup>25</sup> |
|  | Total TB Incidence per 100,000 in 2000 | WHO Global TB report<sup>26</sup> |
|  | Total TB Incidence per 100,000 in 2022 | WHO Global TB report<sup>26</sup> |
|  | TB deaths in HIV-negative people, 2020 | WHO Global TB report<sup>26</sup> |
|  | TB deaths in HIV-negative people, 2022 | WHO Global TB report<sup>26</sup> |
|  | Cumulative notification, 2000 – 2022 | Programmatic data, allowing for +/- 10% uncertainty |
|  | Notification per 100,000 in 2022 | Programmatic data, allowing for +/- 10% uncertainty<sup>26</sup> |
| MDR TB | Multidrug-resistant or rifampicin-resistant TB (MDR/RR-TB) incidence per 100,000, 2015 | WHO Global TB report<sup>26</sup> |
|  | Multidrug-resistant or rifampicin-resistant TB (MDR/RR-TB) incidence per 100,000, 2022 | WHO Global TB report<sup>26</sup> |
|  | Second line treatment initiation in 2022 | Programmatic data, allowing for +/- 10% uncertainty |
| TB HIV | Prevalence of HIV in 2022 | AIDS In UNAIDS<sup>27</sup> |
|  | TB incidence in people living with HIV, 2022 | WHO Global TB report<sup>26</sup> |
|  | Proportion of PLHIV on Antiretroviral therapy in 2022 | From Spectrum AIM<sup>16</sup> |
|  | TB deaths in people with HIV, 2022 | WHO Global TB report<sup>26</sup> |

### Interventions

The model focusses on three major areas of interventions: case detection, treatment and prevention. The intensity of each intervention is guided by the current TB Global Plan or by the country specific National Strategic Plan (NSP). It is assumed that all interventions begin in a designated year and are scaled up linearly over a defined period. This implementation timeline is flexible and can be tailored to reflect global strategies and country-specific priorities and needs.

The model can analyse a range of general interventions, including case detection, various elements of the TB treatment cascade, TB prevention and vaccines. Two example scenarios are illustrated here: 1. the Global Plan intervention scenario for Indonesia (as set out in Global Plan 2023-2030 report) and 2. the NSP scenario for Nigeria (as set by Nigeria’s NSP working group).

#### Global Plan scenario for Indonesia

In the Global Plan intervention scenario, intervention intensities are set at levels sufficient to enable the country to achieve the End TB goals by 2030.^1^ The interventions are scaled up in a linear fashion from 2023 through 2026 and maintained thereafter. For Indonesia, the specific interventions include:

##### Public–private mix (PPM)

PPM efforts are scaled up, assuming an intervention that engages effectively with 80% of private health care providers who are already involved in managing TB, improving their standards of TB diagnosis and TB treatment outcomes to the same level as in the public sector. By improving standards of diagnosis among private providers, the intervention reduces missed opportunities for diagnosis and thus reduces the diagnostic delay. By improving treatment completion, the intervention reduces rates of post-treatment relapse, which would otherwise arise from suboptimal implementation of treatment.

##### Improved routine TB services

TB diagnostics are modernized throughout routine services, i.e., comprehensive replacement of any microscopy-based and clinical diagnosis with rapid molecular tests for TB. In Indonesia, the model assumed that comprehensive use of these diagnostic tools would facilitate recognition of RR status at the point of TB diagnosis.

##### Improved treatment outcomes for Rifampicin Resistant (RR)-TB

All current second-line treatments are replaced with new shorter-term regimens, such that the proportion of treatment success increases to 85%.

##### Upstream case-finding (symptomatic TB)

All activities designed to diagnose symptomatic TB more rapidly than an individual’s first attempt at care seeking. These activities could include active case finding in the community or in healthcare facilities, and also measures such as demand generation, i.e., encouraging those with symptoms to come forward for care more rapidly than they do at present. The model assumed that, together, these measures would reduce the delay-to-diagnosis by 30% for symptomatic individuals.

##### Detecting asymptomatic TB

Measures are in place to find and treat 20% of individuals with asymptomatic TB before they develop symptoms. Note that this intervention is only an example of additional intervention efforts over and above the use of existing tools, which could contribute to meeting the End TB goals. (Alternative strategies might include, for example, new regimens or therapeutic vaccines to reduce post-treatment recurrence.)

##### TB preventive therapy for key and vulnerable populations

There is full uptake of TPT among key and vulnerable populations identified in WHO recommendations, i.e., PLHIV and all-age, close contacts of persons diagnosed with TB.

##### New TB vaccine

A new post-exposure vaccine with 60% efficacy in reducing TB incidence among those with TB infection and conferring immunity for 10 years is rolled out to reach a given coverage of the population (with coverage dependent on the country setting, to meet the End TB goals by 2030). The analysis assumed that a vaccine could be licenced by 2025 and rolled out over the subsequent three years.

#### NSP scenario for Nigeria

As an example, we use the model to assess the impact of the Nigeria NSP 2021-2026 on the country’s tuberculosis (TB) epidemic.^28^

##### Improved notification

To reach the notification target as per the NSP 2021-2026, the following interventions are modelled.

###### i. Public-Private Mix

Scale-up of PPM efforts, assuming an intervention that engages effectively with 35% of private healthcare providers who are already involved in managing TB, improving their standards of TB diagnosis and TB treatment outcomes to the same level as in the public sector. By improving standards of diagnosis amongst private providers, the intervention reduces missed opportunities for diagnosis and thus reduces the diagnostic delay. By improving treatment completion, the intervention reduces rates of post-treatment relapses that would otherwise arise from suboptimal implementation of treatment.

###### ii. Improved routine TB services

Modernization of TB diagnostics throughout routine services, i.e. comprehensive replacement of any microscopy-based and clinical diagnosis with rapid molecular tests for TB. The mode assumed that comprehensive use of these diagnostic tools would improve TB diagnosis to 95% from the current level.

###### iii. Upstream case-finding

All activities designed to diagnose symptomatic and asymptomatic TB more rapidly than an individual’s first attempt at care seeking: these activities could include active case-finding in the community, can also include measures such as demand generation, i.e. encouraging those with symptoms to come forward for care more rapidly than they do at present. The model assumes that together, these measures would reduce the delay-to-diagnosis from the current average of six months to five months.

##### Improved treatment outcomes

The NSP aims for a 92% treatment success rate among drug-susceptible TB cases and seeks to reduce the initial loss to follow-up (LTFU) to less than 5%. This goal is based on lowering the default rate by 5% in the model.

##### Notification among rifampicin-resistant TB

Upfront drug susceptibility testing has been expanded for diagnosed TB patients to promptly identify drug-resistant TB and start second-line treatment. This intervention is scaled up to achieve a 75% case detection rate for DR-TB by 2026, in line with NSP targets.

##### TB preventive treatment, risk groups

Uptake of preventive treatment amongst risk groups identified in WHO recommendations, i.e. people living with HIV and all-age, close contacts of diagnosed TB patients. The coverage among these groups is as per NSP.

## Results

### Calibration

The results of model calibration for Indonesia and Nigeria are shown in Figure 3. This shows the resulting comparisons between model outputs and the eleven indicators as depicted in table 1.

**Figure 3.**
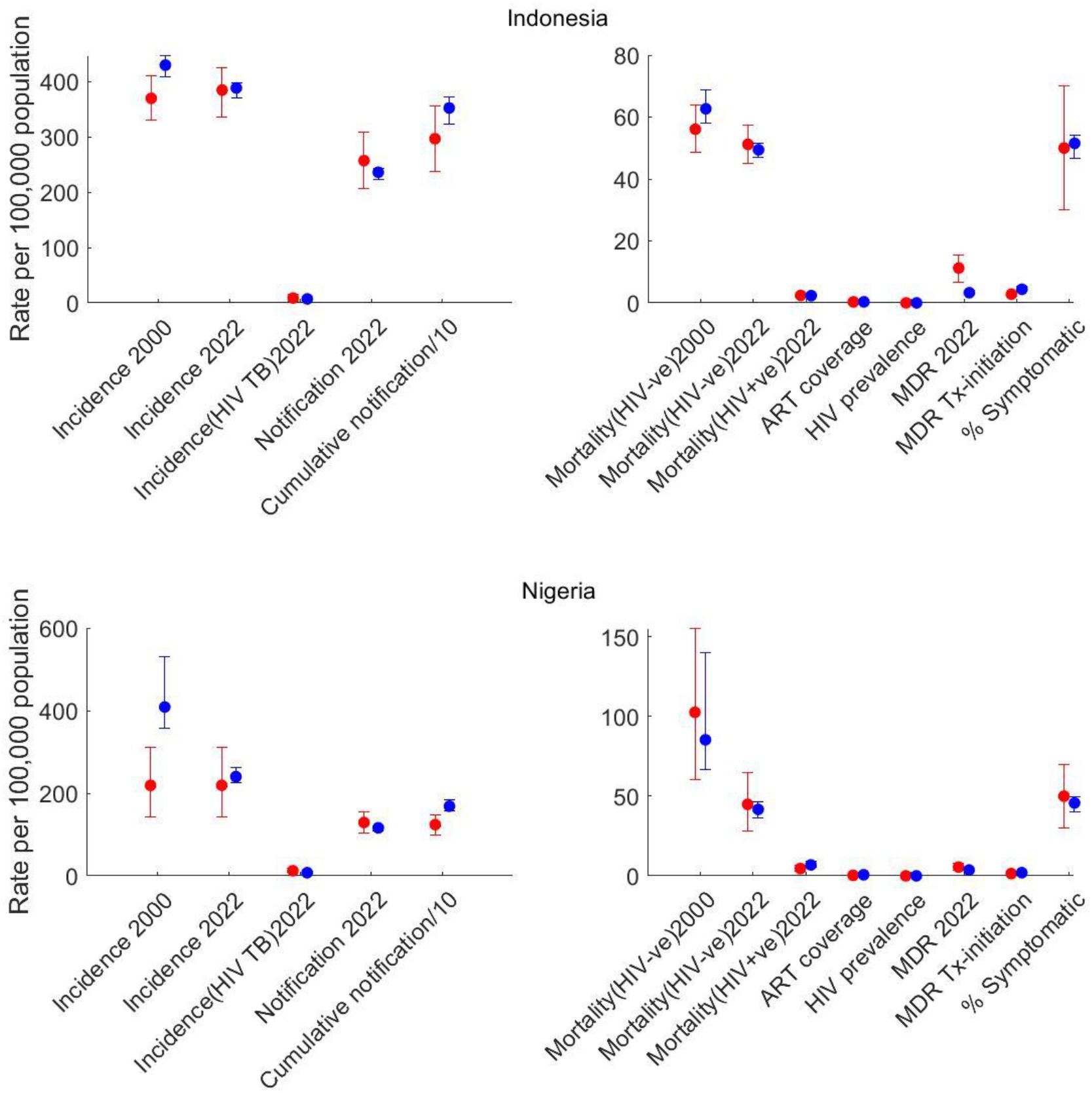
Basic calibration results for Indonesia and Nigeria. Dots show central estimates, while error bars show 95% uncertainty intervals. Here, red markers indicate the data and blue model projected outcomes. The model was unable to fully replicate Nigeria’s 2000 incidence data point, as it predicts a decline in incidence and mortality linked to increased case detection, whereas WHO data indicates a relatively stable incidence from 2000 to 2022.

### Global Plan scenario (Indonesia)

The model projects that Indonesia could achieve the End TB targets set for 2030 by implementing the comprehensive strategy outlined in the Global Plan.^13^ All non-vaccine interventions are scaled up linearly between 2023 and 2026 and subsequently maintained at those levels. The vaccine intervention is assumed to commence in 2028. Figure 4 presents model-based projections of annual TB incidence and mortality rates under various intervention scenarios. In the absence of any additional interventions beyond current programmatic activities, the projected trends in incidence and mortality are shown as the baseline (red lines).

**Figure 4.**
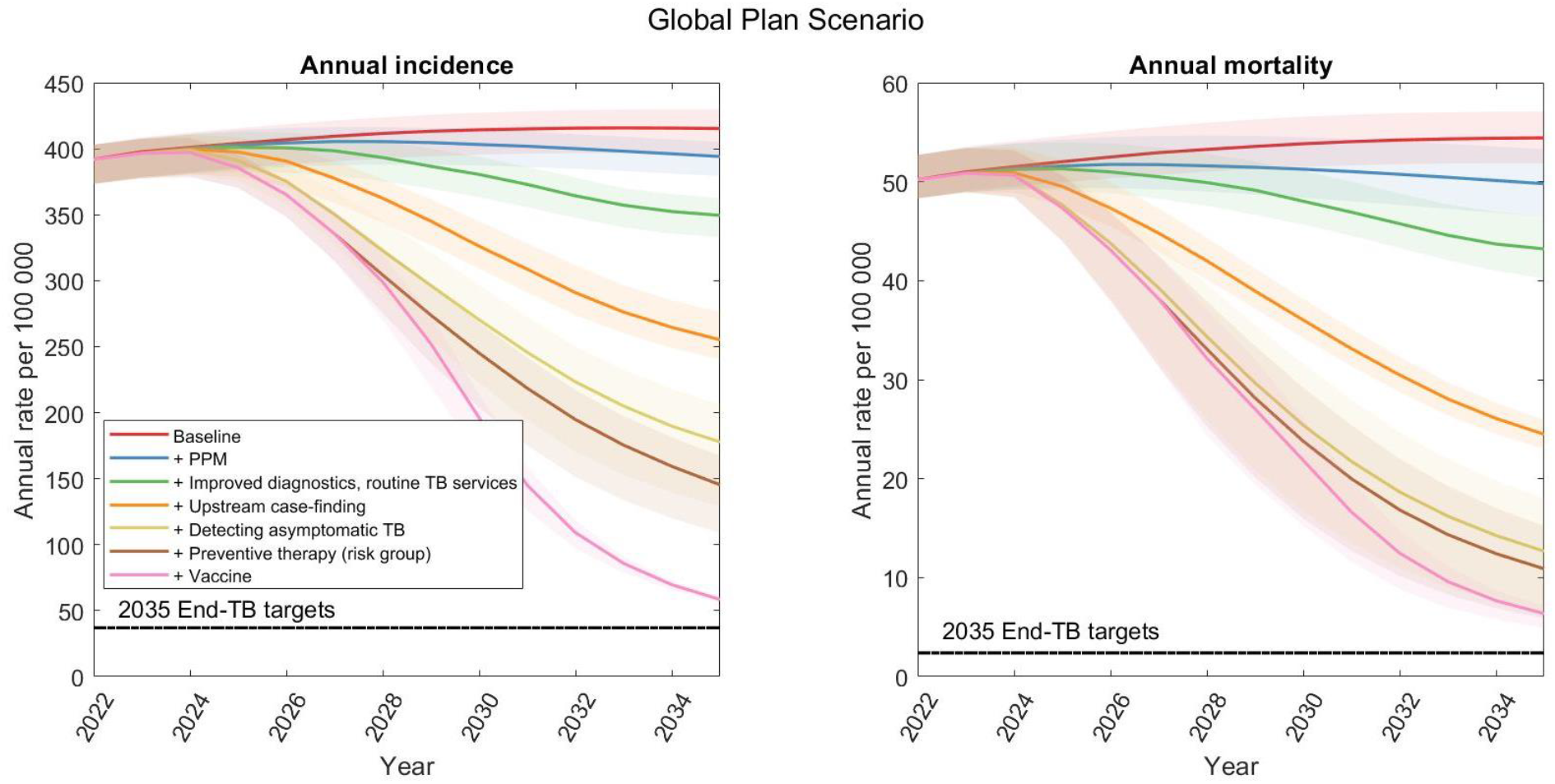
Epidemic dynamics under different intervention scenarios in Indonesia. Shaded regions show 95 per cent credible intervals (CrI), arising from uncertainty in input parameters and in potential future background trends in TB burden. The horizontal, dashed lines show the 2030 targets for incidence (left-hand panel) and mortality (right-hand panel).

The sequential addition of interventions leads to substantial reductions in both incidence and mortality. For instance, implementation of the PPM with improved diagnosis and routine TB service can results in a 12% (CrI: 0.34–23) reduction in incidence and a 18% (CrI: 2.4–32.2) reduction in annual mortality relative to 2023 levels. A combined scale-up of all interventions except the vaccine yields further declines - reducing incidence by 73% (CrI: 61 –85) and mortality by 90% (CrI: 75 – 99) - but remains insufficient to meet the End TB targets by 2030.

To bridge this gap, the model assumes the introduction of a post-exposure vaccine with 60% efficacy at the population level beginning in 2026, with rollout continuing over five years. The inclusion of this vaccine intervention enables Indonesia to reach the 2030 End TB targets, according to model projections.

### Country Scenario (Nigeria)

Figure 5 illustrates the projected impact of the full-scale implementation of the NSP in Nigeria. Each curve represents the impact of individual interventions included in the NSP strategy. The greatest reduction in TB burden is achieved when all interventions are implemented concurrently, as indicated by the magenta line. Comprehensive implementation of the NSP is projected to substantially reduce both TB incidence and mortality, contributing meaningfully toward achieving the End TB targets. In the absence of vaccination, the NSP interventions alone are estimated to reduce TB incidence by 27% and mortality by 37% by 2030, relative to 2022 levels.

**Figure 5.**
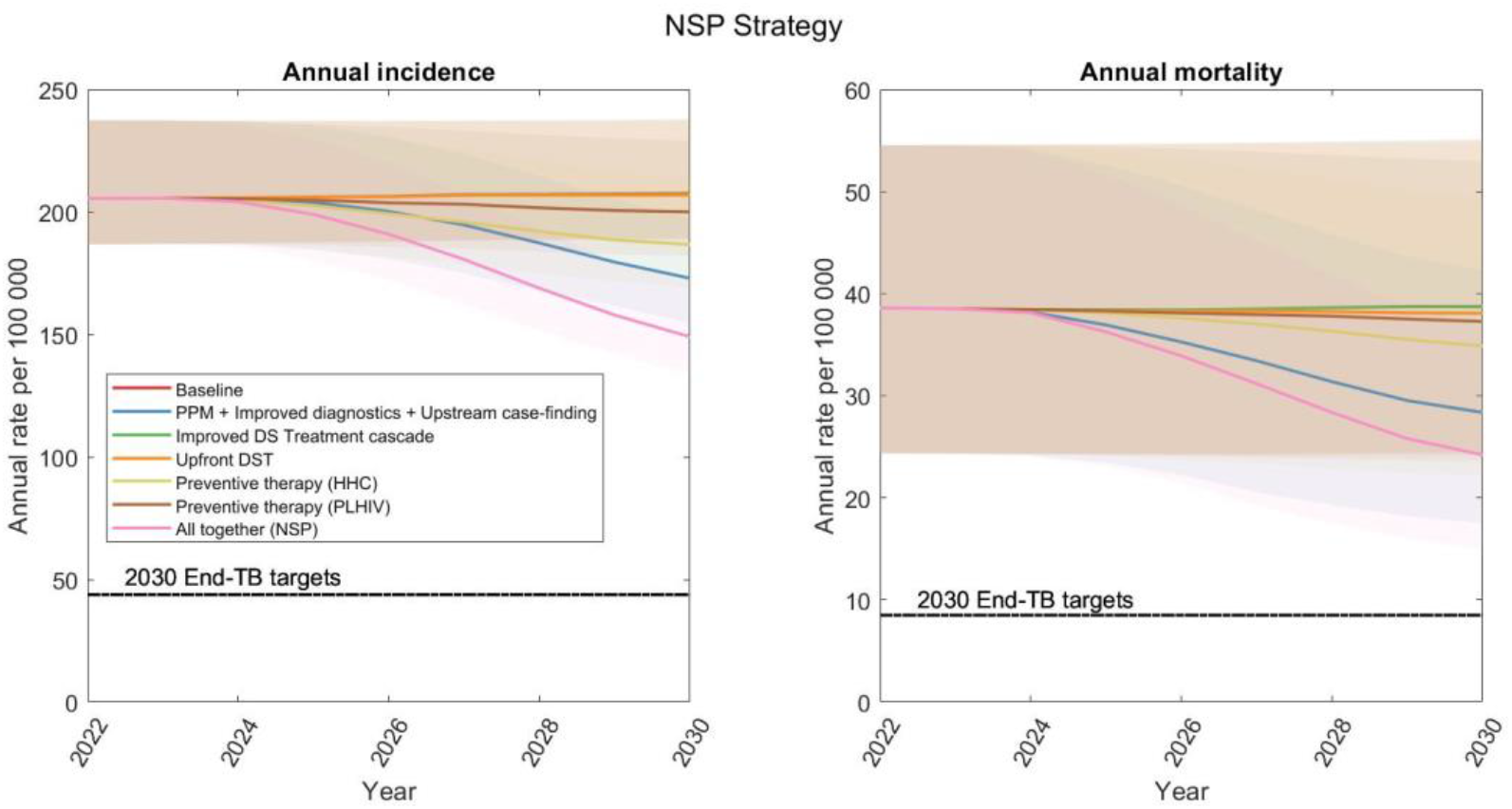
Impact of interventions on the incidence and mortality in Nigeria from 2022 to 2030 under different intervention targets stated in the proposed NSP. ‘PPM + Improved diagnosis + Upstream case finding’ represent a combined effort to reach the stated notification target.

### Lives-saved analysis and primary health care utilization (Indonesia)

The current model can also be used to estimate the number of lives saved by the programs up to 2022, starting from 2000 and 2003, under two scenarios respectively. In the first scenario, outcomes are compared to a hypothetical situation where all public sector TB services ceased in 2000 or the country-specific year of first programmatic results following Global Fund investment, which is 2003 in the case of Indonesia. In the second, TB services are assumed to have continued at their 2000 or 2003 levels without improvement. In both cases, projected deaths under these scenarios are compared to the baseline, and the difference represents the estimated lives saved through the program’s implementation, as shown in Figure 6. These scenarios provide input to return of historic investment analysis as part of the Invest Case for the 8^th^ replenishment and for estimating savings in primary health care utilization. ^29^

**Figure 6.**
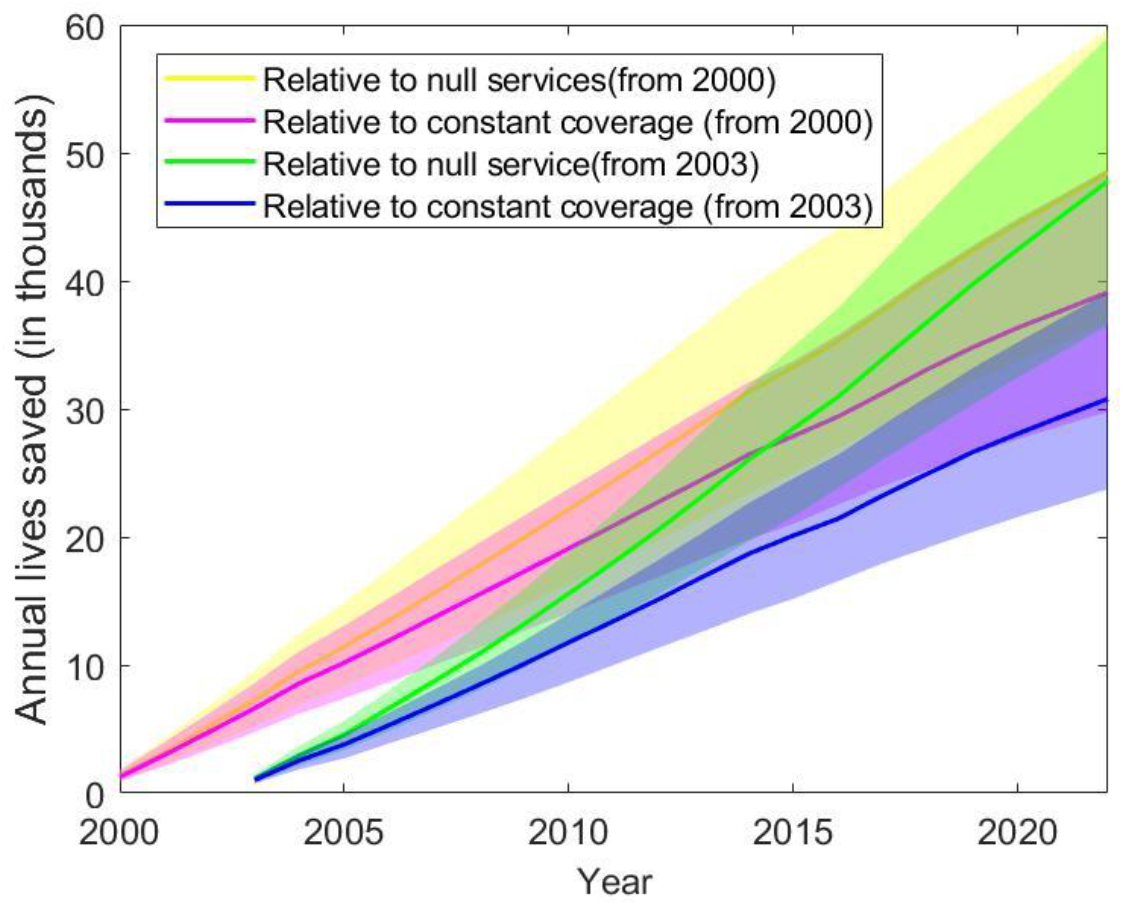
Cumulative lives saved by the TB programmes in Indonesia compared to null service scenario and constant coverage scenario.

## Discussion

This study presents a novel, enhanced global TB dynamical model designed to overcome key limitations of existing frameworks like TIME Impact,^15^ providing a flexible, high-fidelity tool for evaluating national and global tuberculosis control strategies. By capturing critical epidemiological heterogeneities - including asymptomatic TB, age structure, HIV and ART status, and public-private care dynamics - the model offers a more nuanced and policy-relevant assessment of potential intervention scenarios.

Model calibration results demonstrate strong alignment with historical epidemiological data for high-burden countries such as Indonesia and Nigeria (example countries), validating the model’s structure and parameterization. For Indonesia, our findings suggest that a comprehensive scale-up of Global Plan interventions, including enhanced case detection, treatment improvements, and the introduction of a post-exposure TB vaccine, is essential to achieve End TB targets by 2030. Notably, even aggressive implementation of non-vaccine strategies falls short, underscoring the critical role that novel prevention tools, such as vaccines, must play in the final stages of TB elimination.

In contrast, the analysis of Nigeria’s NSP illustrates the considerable gains possible through well-aligned, nationally tailored interventions, even in the absence of a vaccine. While the NSP alone may not suffice to reach End TB targets, it is projected to reduce TB incidence and mortality by 27% and 37%, respectively, by 2030. These gains highlight the value of coherent national planning and the importance of country-level strategic implementation that engages both public and private sectors.

A Target Population (TP) component of the global TB portfolio model was developed to interface between the new global costing and the impact models, and to ensure a good alignment between them. The TP component estimates the various target population needed for costing the algorithms for approximately 20 patient population-based algorithms. These populations are described in detail in a separate paper on the costing model developed together with the dynamical. TB detection, prevention and treatment variables are then aggregated over these populations and used to adjust impact related variables in the global dynamical model. This approach allows for the modelling of cost and impact of strategies that aligns closely with WHO guidelines for the same patient populations – a process that was not possible with existing tools.

A key strength of the model lies in its open-source design which can be accessed through the GitHub repository enabling transparency, reproducibility, and adaptability to country-specific epidemiology and programmatic structures.^30^ Its integration of both diagnostic and treatment pathways across sectors, explicit representation of care-seeking behaviour, and ability to simulate upstream and asymptomatic case finding interventions significantly enhance its realism and policy utility. Moreover, its ability to model vaccine roll-out dynamics, including waning immunity and efficacy assumptions, makes it a critical tool for evaluating the anticipated introduction of TB vaccines.

These functionalities make the model ideally suited for supporting donor decisions, based on investment outcomes such as those presented in the Investment Case for the 8^th^ Global Fund replenishment. The model is also used for studies related to the country-level implementation of the TB Global Plan 2023-2030. Its design allows expansion to study the role of TB determinants, and a study into the cost and impact of global nutrition interventions have been conducted with an adaptation of the model.

The model has potential for country-level applications. The TP component’s close alignment with the TB Target Population component of the WHO’s IHT allows for the model to be used for planning and decision-making at country level through its inclusion as an impact modelling tool in IHT TB, which has recently been completed. Used in this way, the interface of the IHT’s Target Population component serves as a platform for user inputs to the model allowing much closer alignment to country-level contexts than is generally attained when applying the model towards studies focussed on global outcomes.

However, several limitations must be acknowledged. First, while the model accounts for age-dependent mixing patterns, it omits other important mixing variables including, gender, risk and geographical aspects of mixing. Further, while the model captures major determinants of TB infection transmission and control (such as HIV as a risk factor for TB), it currently excludes gender-specific differences in TB burden, comorbidities like undernutrition and diabetes and pulmonary versus extrapulmonary TB distinctions. These omissions, although justifiable for the current scope, may obscure important epidemiological subtleties. Additionally, while HIV is included through stratification, full dynamic modelling of HIV transmission and ART scale-up remains outside the model’s scope. Future iterations could benefit from linking to dynamic HIV models to capture these interactions more comprehensively.

Another key challenge is the reliance on generalized assumptions for certain inputs in the absence of country-specific data - particularly for parameters like treatment adherence and private sector care quality. Although wide priors and Bayesian calibration help account for this uncertainty, localized data collection efforts would greatly improve model fidelity. Finally, while the model runs efficiently for global applications, large-scale scenario testing across many countries still demands considerable computing resources, which may be a barrier for some users.

## Conclusions

In conclusion, this global TB model represents a significant step forward in producing model-based information for strategic TB programme planning. It provides a robust analytical foundation to assess the epidemiological impact of diverse interventions, prioritize investments, and guide policy toward meeting the End TB targets. As funding constraints continue to shape global health priorities, such modelling tools are indispensable for ensuring that limited resources are allocated to the most effective, evidence-based strategies.

## Supporting information

Technical Appendix: Model details

Technical Appendix: Coding guidelines

## Data Availability

All data produced in the present study are available upon reasonable request to the authors

https://github.com/CarelPretorius/GlobalTBTransmissionModel

## Acknowledgements

The authors acknowledge the global portfolio model advisory group convened by TB Modelling and Analysis Consortium (TBMAC), with representatives from various organizations, including experts linked to TBMAC, WHO, STB and the Global Plan’s working group, Global Fund’s modelling guidance group and others whose inputs on the model design, functionality and validation have greatly helped to improve the transmission model component of the global portfolio TB model.

