## Supplementary material for "The *global TB portfolio model*: a tool for projecting the epidemiological impact of TB policy options": Technical Appendix: Model details

### Supplementary documents

Appendix ........ **Error! Bookmark not defined.**

#### Model structure

Figure S1 gives a schematic illustration of the overall model structure, with all model parameters listed in Table S1. The model is stratified by HIV-status, Drug-susceptibility status and by Age groups and vaccination status. The compartment for second line treatment, in case of drug resistance TB is also considered in the model but not shown in the diagram for clarity.

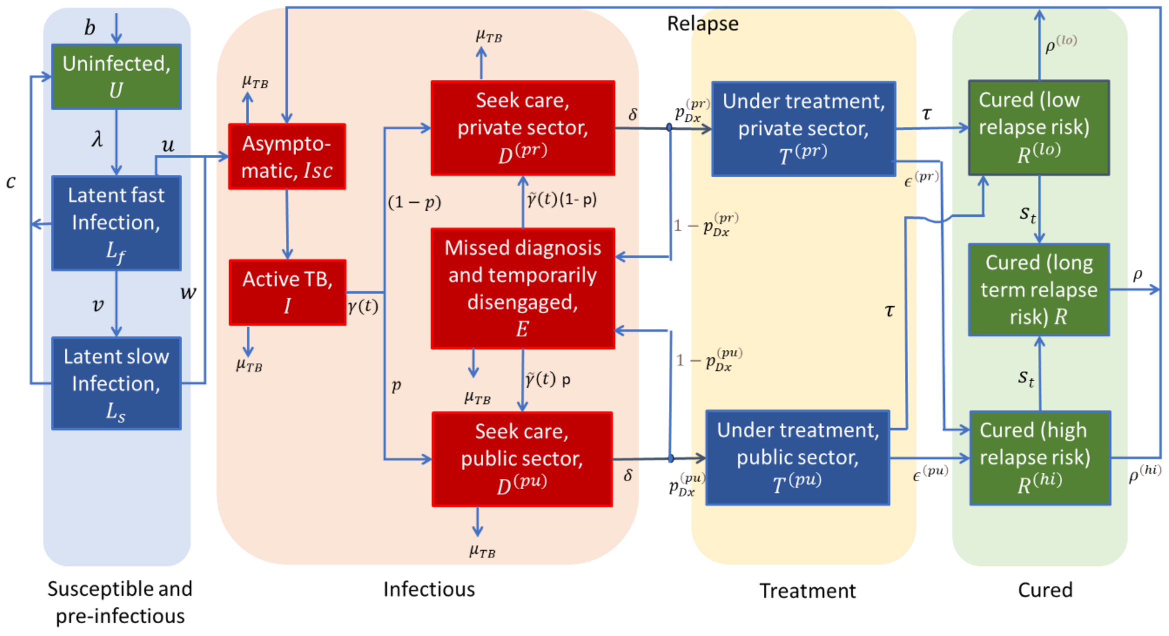

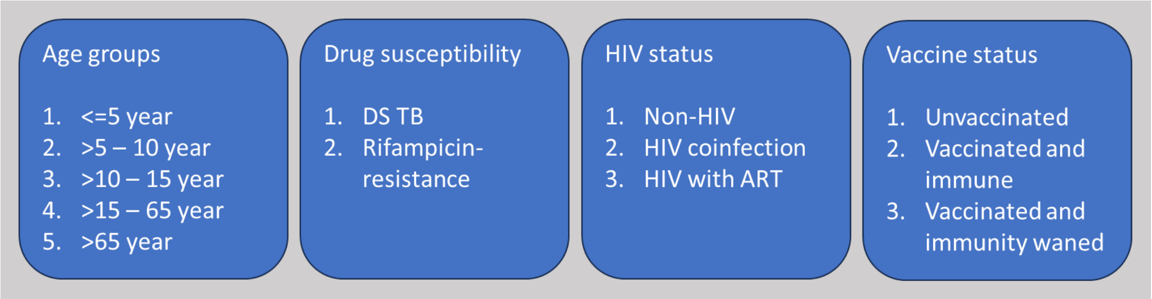

**Figure S1. Schematic illustration of the model structure.** Infectious compartments contributing the force-of-infection are shown in red. For clarity, the diagram omits certain rates incorporated in the model, including self-cure; exogenous reinfection; and background mortality.

#### Governing equations

The governing equations of the model are as follows. All state variables are written as proportions of the population (not as absolute numbers). All variables are divided into five age groups as described in the main text, ($a$ = 0,1,2,3,4) but for clarity we have presented the variable $X_{h,s,a}$ as $X_{h,s}$. And the aging rate from one age group to the next age group is not shown in the set of equations. For example, the rate of aging from $a$ to $a$+1 (where $a$ = 0, 1) is $a_g(a)$ is not shown in the equations. As per HIV status all variables are divided into three categories with indices (h = 0, 1, 2) as: non-HIV ($h_0$.), HIV coinfection ($h_1$) and HIV coinfected individuals with ART ($h_2$). As per drug-susceptibility status all variables are divided into drug susceptible (DS) or drug-resistance (RR) indicated by the indices (s = 0, 1). Based on vaccination status, all variables are categorized as follows: unvaccinated (v = 0), vaccinated and immune (v = 1), and vaccinated with waned immunity (v = 2). For clarity, the index v is not explicitly shown with each variable.

Uninfected ($U_h$):

$$\frac{dU_h}{dt} = b + c\sum_s \left(L_{h,s}^{(fast)} + L_{h,s}^{(slow)}\right) - U_h \epsilon_h \sum_s \lambda_s - \mu_h U_h + f(U_h)$$

for a birth-rate $b$; force-of-infection $\lambda_s$; s denotes the infecting strain (denoting drug-susceptible (s = 0) and drug-resistant (s = 1) TB); h indicates HIV status (denoting HIV negative (h = 0), HIV positive (h = 1) and HIV positive with ART (h = 2); rate of clearance of LTBI $c$; and background mortality rate $\mu_h$. $\epsilon_h$is the relative infectiousness of HIV-coinfected individuals for TB.

Latent, 'fast' infection ($L_{h,s}^{(fast)}$):

$$\frac{dL_{h,s}^{(fast)}}{dt} = \lambda_s \left[U_h + (1-h)\left(R_{h,s}^{(hi)} + R_{h,s}^{(lo)} + R_{h,s}^{(st)}\right)\right] - (\mu_h + u_h + v_h + c)L_{h,s}^{(fast)} + f\left(L_{h,s}^{(fast)}\right)$$

for a progression rate $u_h$; a 'stabilisation' rate (to latent 'slow' status) $v_h$; and protection from reinfection $h$, amongst those previously infected.

Latent, 'slow' infection ($L_{h,s}^{(slow)}$):

$$\frac{dL_{h,s}^{(slow)}}{dt} = v_h L_{h,s}^{(fast)} - (\mu_h + c + w_h)L_{h,s}^{(slow)} + f\left(L_{h,s}^{(slow)}\right)$$

for a reactivation rate $w_h$.

Asymptomatic/Subclinical TB ($I_{h,s}^{(sc)}$):

$$\frac{dI_{h,s}^{(sc)}}{dt} = u_h L_{h,s}^{(fast)} + w_h L_{h,s}^{(slow)} + \rho^{(hi)} R_{h,s}^{(hi)} + \rho^{(lo)} R_{h,s}^{(lo)} + \rho^{(st)} R_{h,s}^{(st)} - \left(\mu_h + \sigma_h + r + a(t)\right)I_{h,s}^{(sc)} + f\left(I_{h,s}^{(sc)}\right)$$

for relapse rates $\rho^{(hi)}, \rho^{(lo)}, \rho^{(st)}$ ; rate of developing clinical symptom $r$ and self-cure rate $\sigma_h$. The term $a(t)$ represents the time-dependent effect of active case-finding.

Clinical TB ($I_{h,s}$):

$$\frac{dI_{h,s}}{dt} = r\, I_{h,s}^{(sc)} - (\gamma + \mu_h^{(TB)} + \sigma_h + a(t))I_{h,s} + f\left(I_{h,s}\right)$$

for care-seeking rate $\gamma$ and TB mortality rate $\mu_h^{(TB)}$.

Presented for diagnosis with provider type $s$, ($D_{h,s}^{(s)}$):

$$\frac{dD_{h,s}^{(pu)}}{dt} = p\,(\gamma\, I_{h,s} + \tilde{\gamma}\, E_{h,s}) - \left(\mu_h^{(TB)} + \sigma_h + \delta\right)D_{h,s}^{(pu)} + f\left(D_{h,s}^{(pu)}\right)$$

$$\frac{dD_{h,s}^{(pr)}}{dt} = (1-p)\,(\gamma\, I_{h,s} + \tilde{\gamma}\, E_{h,s}) - \left(\mu_h^{(TB)} + \sigma_h + \delta\right)D_{h,s}^{(pr)} + f\left(D_{h,s}^{(pr)}\right)$$

Here, $p$ and $(1-p)$ are, respectively, the proportion-of-presentation to healthcare providers in the public and private sectors. Rate-of-offering diagnosis $\delta$. Here, $\tilde{\gamma}$ is analogous to $\gamma$ but attached to individuals who remain undiagnosed despite having previously sought care (i.e. compartment $E_h$, below).

On TB treatment with provider type $po$, with DS TB ($T_{h,0}^{(po)}$) initiating first line treatment:

$$\frac{dT_{h,0}^{(pu)}}{dt} = \delta\, p_{Dx}^{(pu)} \left(D_{h,0}^{(pu)} + \left(1 - DST_{(+ve)}\right)D_{h,1}^{(pu)}\right) + a(t)(I_{h,0} + I_{h,0}^{(sc)}) - \left(\mu_h + \tau + \epsilon^{(pu)} + r_{aqr}\right)T_{h,0}^{(pu)} + f\left(T_{h,0}^{(pu)}\right)$$

$$\frac{dT_{h,0}^{(pr)}}{dt} = \delta\, p_{Dx}^{(pr)} D_{h,0}^{(pr)} - \left(\mu_h + \tau + \epsilon^{(pr)} + r_{aqr}\right)T_{h,0}^{(pr)} + f\left(T_{h,0}^{(pr)}\right)$$

for a treatment completion rate $\tau$; and a treatment interruption rate $\epsilon^{(po)}$. $a(t)$ is the rate of active case finding. $r_{aqr}$is the rate of acquisition MDR during treatment and they move to $T_{h,1}^{(pu)}$and $T_{h,1}^{(pr)}$ respectively and not shown in the equations.

Drug resistance TB initiating second line treatment ($S_{h,1}^{(pu)}$) :

$$\frac{dS_{h,1}^{(pu)}}{dt} = \delta\, p_{Dx}^{(pu)}\, DST_{(+ve)}\, D_{h,1}^{(pu)} + SLtrans * T_{h,0}^{(pu)} - \left(\mu_h + T + \epsilon 2^{(pu)}\right)T_{h,0}^{(pu)} + f\left(S_{h,1}^{(pu)}\right)$$

For treatment completion rate $T$; and a treatment interruption rate $\epsilon 2^{(po)}$. SLtrans represents transfer to SL treatment during FL treatment.

Missed diagnosis and temporarily disengaged from care-seeking ($E_{h,s}$):

$$\frac{dE_{h,s}}{dt} = \delta\left(1 - p_{Dx}^{(pu)}\right) D_{h,s}^{(pu)} + \delta\left(1 - p_{Dx}^{(pr)}\right) D_{h,s}^{(pr)} - \left(\mu_h^{(TB)} + \sigma_h + \tilde{\gamma}\right) E_{h,s} + f(E_{h,s})$$

Recovered with low relapse risk, following treatment completion ($R_h^{(lo)}$):

$$\frac{dR_{h,s}^{(lo)}}{dt} = \tau\left(T_{h,s}^{(pu)} + T_{h,s}^{(pr)}\right) - [(1-h)\lambda_s + \rho^{(lo)} + \mu_h + s_t]R_{h,s}^{(lo)} + f\left(R_{h,s}^{(lo)}\right)$$

for a rate of 'stabilisation' of relapse risk $s_t$.

Recovered with high relapse risk, following treatment completion ($R_{h,s}^{(hi)}$):

$$\frac{dR_{h,s}^{(hi)}}{dt} = \left(\epsilon^{(pu)}\, T_{h,s}^{(pu)} + \epsilon^{(pr)}\, T_{h,s}^{(pr)}\right) - [(1-h)\sum_s \lambda_s + \rho^{(hi)} + \mu_h + s_t]R_{h,s}^{(hi)} + f\left(R_{h,s}^{(hi)}\right)$$

Long-term, 'stabilised' relapse risk ($R_{h,s}^{(st)}$):

$$\frac{dR_{h,s}^{(st)}}{dt} = s_t\left(R_{h,s}^{(lo)} + R_{h,s}^{(hi)}\right) - \left[(1-h)\lambda_s + \rho^{(st)} + \mu_h\right]R_{h,s}^{(st)} + f\left(R_{h,s}^{(st)}\right)$$

Force-of-infection ($\lambda_{s,a}$):

$$\lambda_{0,a} = \sum_h \beta_{h,ds}\, C_{ij}\, (k\, I_{h,0}^{(sc)} + I_{h,0} + E_{h,0} + D_{h,0}^{(pu)} + D_{h,0}^{(pr)})$$

$$\lambda_{1,\mathrm{a}} = \sum_h \beta_{h,dr}\, C_{ij}\, (k\, I_{h,1}^{(sc)} + I_{h,1} + E_{h,1} + D_{h,1}^{(pu)} + D_{h,1}^{(pr)})$$

Where $\beta_{h,ds}$ is the rate-of-transmission associated with drug-susceptible and $\beta_{h,dr}$ is associated with drug-resistance TB disease. Both are accompanying with HIV status. HIV-positive TB can be less infectious than HIV-negative TB, $\beta_{1,s}$is expected to be lower in value than $\beta_{0,s}$ and $\beta_{2,s}$. Accordingly, it was assumed that $\beta_{0,s}$ = $\beta_{2,s}$= $m\ \beta_{1,s}$, for a parameter $m$ to be calibrated, and constrained to be between 0 and 1. Therefore, $\epsilon_h$= [1, m, 1]. $k$ is the infectiousness of subclinical TB relative to clinical TB. $C_{ij}$ is the contact matrix between different age groups.

In all the above equations, $f(.)$ denotes transitions between HIV states. For any given state variable $X_h$, it is defined as follows:

$$f(X_h) = \begin{cases} -r_{HIV}X_0, & if\ h = 0 \\ r_{HIV}X_0 - r_{ART}X_1, & if\ h = 1 \\ r_{ART}X_1, & if\ h = 2 \end{cases}$$

where $r_{HIV}$ denotes the per-capita rate of acquiring HIV, and $r_{ART}$ denotes the per-capita rate of initiating ART.

#### Age structure and aging

The age structure of the model is depicted below. The age categories ≤5 years, >5–10 years, >10–15 years, >15–65 years, and >65 years are chosen to inform WHO recommended screening and diagnostic algorithms for specific age groups, with the 65+ category added to capture the high mortality of this age group.

The aging rates are based on input from the Spectrum IHT demographical model (DemProj) which uses WPP data set up its demographical structure.[1] The rate at which individuals age out of age category with age bounds [a1,b1] (e.g. 0-4 years), and into the next age category [a2,b2] (5-9 years) is given by pop[b1]/(pop[a1]+...pop[b1]).

These rates are prepared for each country, by year, with calls to the Spectrum DemProj model allowing the global TB model to maintain an approximate age structure for a given population.

Since the risk of contracting directly transmitted infections depends on interaction patterns between individuals, mathematical models often employ contact matrices to describe the spread of infectious pathogens. Age-structured models typically represent the interaction rates between age groups using a mixing matrix, where each element indicates the frequency of contact between individuals from specific subgroups (such as age groups), as defined by the corresponding rows and columns. We adjusted the force of infection using country-specific age-contact matrices, based on the published work of Prem et al., to capture interactions across different age groups.[2]

#### Model calibration

The model has been calibrated with the country specific data as described in table 1 in the main text. We sampled from the posterior density using adaptive Bayesian Markov Chain Monte Carlo simulation as described in the Calibration section of the main article.[3] Figure S2 below shows the trace arising from the MCMC calibration for Indonesia.

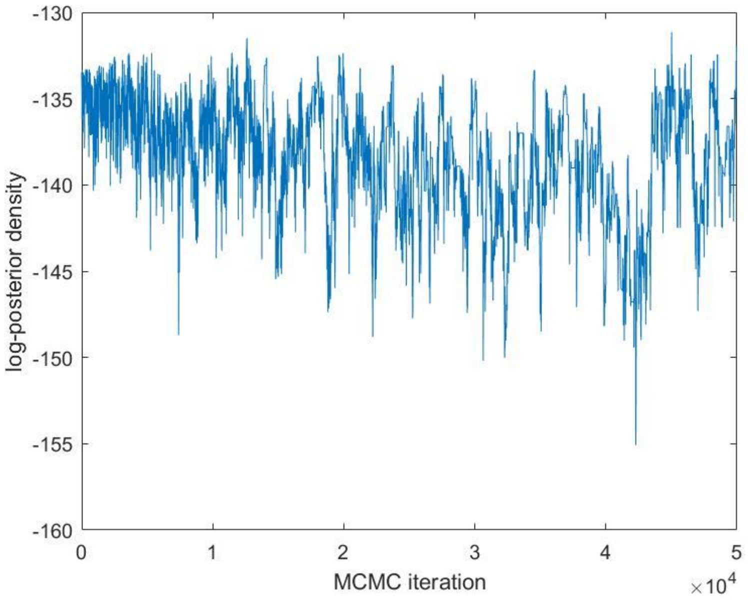

Figure S2. Trace plot arising from MCMC calibration, showing the log-posterior density over 50,000 iterations

#### Parameter values

*Table S1. Table of parameters and their values (For Indonesia and Nigeria)*

| **Parameter** | **Symbol** | **Value (for Indonesia)** | **Value (for Nigeria)** | **Source/Notes** |
|---|---|---|---|---|
| | Natural history | | | |
| Infection rate (number of annual infections per case) | $\beta_{h,ds}$ | 8.6 (7.6 – 9.3) | 10.5 (9 – 13.4) | Model calibration, with priors U[0, 30] |
| Relative infection rate for RR-TB | $rf\beta_{h,dr}$ | 0.66 (0.58 – 0.76) | 0.70 (0.55 – 0.86) | Model calibration |
| Relative infectiousness of HIV-coinfected individuals for TB | $\epsilon_1$ | 0.94 (0.71 – 0.99) | 0.80 (0.03 – 0.99) | Model calibration |
| Rate of developing clinical symptoms | $r$ | 11 (5 – 25) | 29 (20 - 43) | Model calibration |
| Rate of acquired RR TB | $r_{aqr}$ | 0.015 (0.011 – 0.024) | 0.019 (0.005 – 0.03) | Model calibration |
| Per-capita annual rate of progression from 'fast' latent infection | $u_0$<br>$u_1$<br>$u_2$ | 0.0826<br>7.4 (5 – 8)<br>2.9 (2.0 – 3.3) | 0.0826<br>0.2 (0.1 – 0.4)<br>0.08 (0.04 – 0.16) | Menzies (2018)[4] for central value, and U[0.1 - 10] on multiplying factor |
| Per-capita annual rate of reactivation from 'slow' latent infection | $w_0$<br>$w_1$<br>$w_2$ | 0.0006<br>0.05 (0.03 – 0.06)<br>0.02 (0.01 – 0.024) | 0.0006<br>0.0014(0.0007 – 0.0029)<br>0.0006 (0.0003 – 0.0012) | Menzies (2018)[4] for central value, and U[0.1 - 20] on multiplying factor |
| Per-capita annual rate of stabilisation from 'fast' to 'slow' latent status | $v_0$<br>$v_1$<br>$v_2$ | 0.872<br>0<br>0.872 | | Menzies (2018)[5] for central value, and taking uniform priors of +/- 25% |

| | | | | |
|---|---|---|---|---|
| Per-capita annual rate of self-clearance of latent TB | $c$ | 0.031 (0.022 – 0.035) | | Emery (2021)[5] for central value, and taking uniform priors of +/- 25% |
| Per-capita annual rate of TB mortality while untreated among HIV -ve individuals | $\mu_0^{(TB)}$ | 0.27 (0.24 – 0.30) | 0.32(0.30 – 0.33) | Model Calibration with the central value from Tiemersma EW (2011)[6] |
| Per-capita annual rate of HIV mortality | $\mu_1^{(HIV)}$ | 0.05 (0.03 – 0.06) | 0.03 (0.01 – 0.06) | |
| Per-capita annual rate of TB mortality while untreated among HIV +ve with ART individuals | $\mu_2^{(TB)}$ | 0.79 (0.62 – 0.97) | 10 (8 - 13) | |
| Per-capita annual rate of TB self-cure from HIV-ve ($\boldsymbol{h_0}$ )and HIV +ve ($\boldsymbol{h_2}$) with ART individuals | $\sigma$ | 0.17 (0.14 – 0.18) | 0.15 (0.14 – 0.18) | |
| Protection from reinfection amongst those with prior infection | $h$ | 0.8 (for $h_0$)<br>0 (for $h_1$)<br>0.8 (for $h_2$) | | Assumption: 80% protection among those who are HIV-ve, or on ART |
| Per-capita annual rate of relapse in first two years after treatment completion | $\rho^{(lo)}$ | 0.032 (0.03 – 0.04) | | Thomas A et al (2005)[7], Romanowski (2019)[8], Menzies (2009)[9] and Weis (1994)[10] , with uniform prior using intervals of $\pm$ 5% |
| Per-capita annual rate of relapse in first two years after self-cure or incomplete treatment | $\rho^{(hi)}$ | 0.14 (0.11 – 0.17) | | |
| Per-capita annual rate of relapse >two years after last TB episode | $\rho$ | 0.0015 (0.0011 – 0.0018) | | Most relapse occurs in first two years |

| | | | | | |
|---|---|---|---|---|---|
| Per-capita annual rate of 'stabilising' from high to low relapse risk | | $s$ | 0.5 | | after recovery: Guerra-Assuncao (2015)[11] |
| | | TB services | | | |
| Rate-of-presentation to care, first careseeking visit | In, 1997 | $\gamma^{(1997)}$ | 1.77 (1.59 – 1.98) | 1.65 (1.45 – 2.05) | Model calibration, with priors U[0.1, 10] for 2011; and for 2020 taking U[1, 10] as multiplying factor on $\gamma^{(1997)}$ |
| | In, 2022 | $\gamma^{(2022)}$ | 1.8 (1.6 – 2.0) | 2.13 (1.48 – 3.09) | |
| Rate-of-presentation to care, second and subsequent careseeking visits | Assuming no change in 1997 and 2022 | $\tilde{\gamma}$ | 32 (22 – 48) | 2.0 (1.4 – 3.8) | Model calibration, taking U[1, 40] for multiplying factor on $\gamma^{(1997)}$ |
| Probability that a TB patient visits public provider, per careseeking attempt | In, 2022 | $p^{(2022)}$ | 0.5 (0.4 – 0.6) | 0.55 (0.48 – 0.60) | Model calibration |
| Per-capita annual rate of offering diagnosis | | $\delta$ | 52 | 52 | Assumption, corresponding to 1 week |
| Probability of successful TB diagnosis and treatment initiation per careseeking visit | Public sector | $p_{Dx}^{(pu)}$ | 0.86 (0.76 – 0.90) | 0.78 (0.75 – 0.89) | U[0.3, 0.9], motivated by Subbaraman (2016) [12] |
| | Private sector | $p_{Dx}^{(pr)}$ | 0.27 (0.21 – 0.34) | 0.49 (0.43 – 0.62) | U[0.3, 0.9], assumption |
| Proportion of RR TB recognised in 2022 | $DST_{(+ve)}$ | | 0.10 (0.01 – 0.28) | 0.025 (0 – 0.23) | Model calibration, allowed range U[0, 1] |

| | | | | | |
|---|---|---|---|---|---|
| Year of ART initiation | | | 2004 | 2004 | |
| Rate of ART initiation | | $r_{ART}$ | 0.04 (0.03 – 0.05 | 0.13 (0.09 – 0.18) | Calibration |
| Per-capita annual rate of first-line treatment completion (DS TB) | | $\tau$ | 2 | | Corresponds to average duration of 6 months |
| Per-capita annual rate of second-line treatment completion (RR TB) | | $\tau_2$ | 0.5 | | Corresponds to average duration of 24 months |
| Per-capita annual rate of treatment interruption | Public sector | $\epsilon^{(pu)}$ | 0.51 (0.47 – 0.55) | 0.64 (0.62 – 0.66) | Calculated using $\epsilon^{(pu)} = \frac{1-P}{P}\tau$ , for treatment completion rate $P$, and assuming U[0.75, 0.95] for $P$ |
| | Private sector | $\epsilon^{(pr)}$ | 0.63 (0.50 – 1.2) | 1.22 (0.51 – 2.70) | As above, but assuming U[0.4, 0.8] for $P$ |
| | | Demographics | | | |
| Per-capita annual rate of background mortality | | $\mu$ | 1/68 | 1/54 | Corresponds to average lifespan (World Bank 2022) [1] |
| Annual population growth rate in 2022 | | $p_{gth}$ | 0.013 | 0.044 | |
| Rate of aging between successive age groups | | $a_{12}$<br>$a_{23}$<br>$a_{34}$<br>$a_{45}$ | 0.189<br>0.171<br>0.180<br>0.008 | 0.176<br>0.170<br>0.160<br>0.005 | |
| Age-contact matrix | | $C_{ij}$ | $C_{ij}^{IDN}$ (see below) | $C_{ij}^{NGA}$ (see below) | Prem et al[2] |

$$C_{ij}^{IDN} = \begin{pmatrix} 0.27 & 0.16 & 0.07 & 0.59 & 0.03 \\ 0.16 & 0.91 & 0.38 & 0.79 & 0.04 \\ 0.09 & 0.24 & 1.62 & 1.32 & 0.04 \\ 0.89 & 0.86 & 0.93 & 14.9 & 0.35 \\ 0.03 & 0.03 & 0.03 & 0.17 & 0.04 \end{pmatrix}$$

And,

$$C_{ij}^{NGA} = \begin{pmatrix} 0.64 & 0.33 & 0.13 & 0.90 & 0.04 \\ 0.33 & 2.16 & 0.61 & 1.17 & 0.08 \\ 0.16 & 0.39 & 1.65 & 1.54 & 0.09 \\ 0.91 & 0.85 & 0.85 & 11.8 & 0.21 \\ 0.02 & 0.02 & 0.02 & 0.07 & 0.02 \end{pmatrix}$$

**Table S1. List of model parameters, values and sources.** The notation $U[x, y]$ denotes a uniform probability distribution on the range $[x, y]$. Otherwise, parameter values in round brackets show 95% percentiles from the respective marginal posterior densities.

#### Interventions

The model focusses on three major areas of interventions: case detection, treatment and prevention, as shown in Figure S3. The numeric values mentioned here are based on the current TB Global Plan analysis and may need adjustment when numerical targets for total notification, for example, are set in a National Strategic Plan (NSP). The specific values mentioned below are illustrative and show the impact mechanisms of the model.

Note that the flows resulting from a vaccine program are not shown in Figure S3. Vaccination is handled by an additional dimension to the model and works similarly to the TPT flows shown.

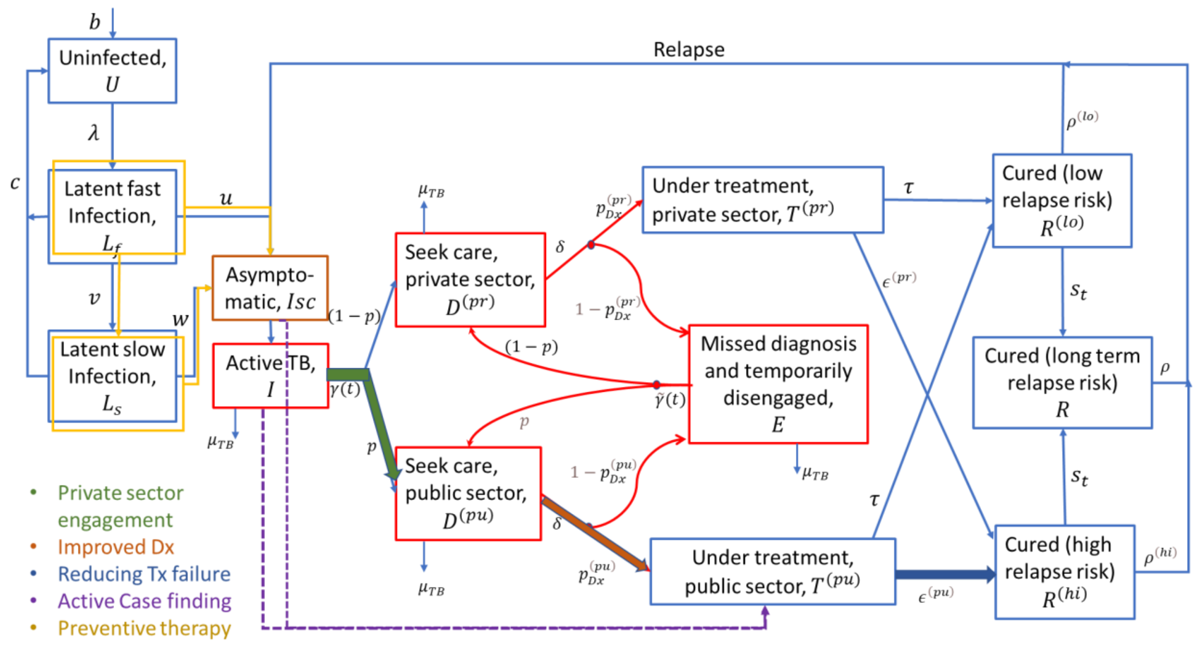

**Figure S3.** ***Schematic illustration of intervention structure.*** *Key intervention shown as slows between model compartments.*

#### Case detection

- Strengthen existing systems:
  - Expand laboratory facilities- to increase uptake of public-sector TB care provision, over 5 years.
    (controlled by changing parameter $\gamma(t)$.)
  - Private sector engagement- recruit additional individuals of private-sector providers, to attain the same quality of care as in the public sector, over 5 years.
    (controlled by changing parameter p)
  - Improved diagnosis for finding DS TB – probability of successful diagnosis per care-seeking visit could be improved by using new tolls.
    (controlled by changing parameter $p_{Dx}^{(pu)}$)
  - Improved diagnosis for finding RR TB - proportion of RR TB recognised could be improved using new tools. (controlled by changing parameter $DST_{(+ve)}$)

- Accelerated Case Detection:
  - Upstream case finding (symptomatic TB): These represent activities that are designed to diagnose symptomatic TB more rapidly than an individual's first attempt at care seeking. These activities could include active case finding in the community, and measures such as demand generation, i.e., encouraging those with symptoms to come forward for care more rapidly than they do at present. By adjusting the one of the rate parameters indicated by violet colours in figure S3.
    (This can be controlled by rate of transition from $I$ to $T_{pu}$)

  - Detecting asymptomatic/subclinical TB: Measures are put in place to find and treat individuals with subclinical TB before they develop symptoms. (by adjusting the one of the rate parameters indicated by violet colours)
    (This can be controlled by rate of transition from $I_{SC}$ to $T_{pu}$)

#### Treatment cascade

- Improved DS treatment success:
  - Successful treatment completion amongst diagnosed cases – this can be obtained by reducing failure and defaults during treatment.
    (controlled by adjusting the parameter $\epsilon^{(pu)}$)

- Improved RR treatment:
  - All current second-line treatments are replaced with new regimens, such that the proportion of treatment success increases to 90%.
    (controlled by adjusting the parameter proportion of success (not depicted in the figure))
  - Shorter duration of second line treatment – this will reduce default during second line treatment. (controlled by adjusting the parameter $\tau_2$)

#### Prevention

- TB preventive therapy:
  - Full uptake of TPT among all household-contacts, PLHIV and all-age and key- vulnerable populations identified in WHO recommendations. Interventions to PLHIV is modelled directly by reducing progression and activation rate to those who are on on ART ($i.e. h_2$).

- Impact of TB preventive therapy among household-contacts and vulnerable population is modelled using the indirect approach described in Mandal et al (2020).[13] (Modelled by reducing the parameter $u_h$, $w_h$ among HIV-negative individuals)

- TB vaccine:
  - The impact of a post-exposure vaccine with a stated efficacy and a target coverage. As of now coverage was chosen in such a way to meet the End TB goals by 2030/2035. (Modelled by adjusting the vaccination rate from v0 to v1).
