## Supplementary material for "The *global TB portfolio model*: a tool for projecting the epidemiological impact of TB policy options": Technical Appendix: Coding guidelines

This document outlines coding guidelines for model code available at the GitHub repository for the countries Indonesia and Nigeria:

https://github.com/CarelPretorius/GlobalTBTransmissionModel

Table 1. List of model parameters and their symbol as depicted in figure S1 and the model equations in the Supplementary document.

| **Parameter** | **Symbol (diagram and equation)** | **Symbol used in the code (see the GitHub link)** |
|---|---|---|
| **Natural history** | | |
| Infection rate (number of annual infections per case) | $\beta$,<br>$\beta_{mdr}$,<br>$\beta_{hiv}$ | $r.beta$<br>$r.beta * p.rfbeta\_mdr$<br>$r.beta * p.rfbeta\_hiv$ |
| Per-capita annual rate of progression from 'fast' latent infection | $u$ | $r.progression$ |
| Per-capita annual rate of stabilisation from 'fast' to 'slow' latent status | $v$ | $r.LTBI\_stabil$ |
| Per-capita annual rate of reactivation from 'slow' latent infection | $w$ | $r.reactivation$ |
| Per-capita annual rate of self-clearance of latent TB | $c$ | $r.LTBI\_cure$ |
| Per-capita annual rate of developing symptoms, amongst subclinical TB | $r$ | $r.sym$ |
| Per-capita annual rate of TB mortality while untreated | $\mu_{TB}$ (general population)<br><br>$\mu_{TB}$ (among HIV) | $r.mort\_TB(1)$<br><br>$r.mort\_TB(2)$ |
| Per-capita annual rate of TB self-cure | $\sigma$ | $r.self\_cure$ |

| | | | |
|---|---|---|---|
| Protection from reinfection amongst those with prior infection | | $h$ | $p.imm$ |
| Per-capita annual rate of relapse in first two years after treatment completion | | $\rho^{(lo)}$ | $r.relapse(1)$ |
| Per-capita annual rate of relapse in first two years after self-cure or incomplete treatment | | $\rho^{(hi)}$ | $r.relapse(2)$ |
| Per-capita annual rate of relapse>two years after last TB episode | | $\rho$ | $r.relapse(3)$ |
| Per-capita annual rate of 'stabilising' from high to low relapse risk | | $s_t$ | $r.s$ |
| Rate-of-presentation to care, first care-seeking visit | In 1997<br>In 2022 | $\gamma(1997)$<br>$\gamma(2022)$ | $r.cs\_1997$<br>$r.cs\_1997 * rf\_cs2022$ |
| Rate-of-presentation to care, second and subsequent care-seeking visits | In 1997<br>In 2022 | $\tilde{\gamma}\ (1997)$<br>$\tilde{\gamma}\ (2022)$ | $r.cs\_1997 * rf\_cs2$<br>$r.cs\_1997 * rf\_cs2$ |
| Probability that a TB patient visits public provider, per care-seeking attempt | In 2022 | $p$ | $p.pu\_2022$ |
| Per-capita rate of offering diagnosis | | $\delta$ | $r.Dx$ |

| | | | |
|---|---|---|---|
| Probability of successful TB diagnosis and treatment initiation per care-seeking visit | Public sector | $p_{Dx}^{(pu)}$ | $p.Dx(1)$ |
| | Private sector | $p_{Dx}^{(pr)}$ | $p.Dx(2)$ |
| Per-capita annual rate of treatment completion | | $\tau$ | $r.Tx$ |
| Per-capita annual rate of treatment interruption | Public sector | $\epsilon^{(pu)}$ Calculated using $\epsilon^{(pu)} = \frac{1-P}{P}\tau$ , for treatment completion rate $P$, and assuming U[0.75, 0.95] for $P$ | $r.default(1)$ |
| | Private sector | $\epsilon^{(pr)}$ | $r.default(2)$ |
| **Demographics** | | | |
| Per-capita annual rate of background mortality | | $\mu$ (general population)<br>$\mu$ (among HIV) | $r.mort(age)$<br>$r.HIV\_mort$ |

### Model implementation

The model is implemented in both Matlab and Python and the code is hosted in a GitHub repository. The sections below define and describe the syntax and functions used:

- Table 2 defines states names and symbols used of compartments and variables in the model.
- Table 3 defines auxiliary measures (outputs) that are needed for fitting and display purposes.
- Table 4 defines the functions that structure the model setup, model calibration and projection steps.
- Table 5 states the natural history parameters and their data values and ranges, with references.
- Table 6 state parameters that are estimated within assigned ranges as part of the fitting process.

- Table 7 state time-dependent variables that are linked to interventions.
- Table 8 states the WHO data used for calibration purpose.

### Compartments and variables

*Table 1: Defining compartments and variables.*

| | | |
|---|---|---|
| gps.vacc | | Grouping by vaccination status |
| | v0 | Unvaccinated |
| | v1 | Vaccinated, immune |
| | v2 | Vaccinated, waned immunity |
| gps.hiv | | Grouping by HIV status |
| | h0 | hiv-negative |
| | h1 | hiv-positive |
| | hart | hiv-positive, with ART |
| gps.strains | | Grouping by drug resistance status |
| | ds | Drug susceptible TB |
| | mdr | Drug-resistant (rifampicin-resistant) TB |
| gps.provs | | Grouping by provider type |
| | pu | NTP provider (public, or notifying) |
| | pr | non-NTP provider (private, or non-notifying) |
| | U | Uninfected |
| | Lf | Latent fast |
| | Ls | Latent slow |
| | Isc | Subclinical infection |
| | I | Infection with clinical symptoms |
| | E | Missed diagnosis and temporarily disengaged from care-seeking |
| | Rlo | Recovered with low relapse risk, following treatment completion |

| | |
|---|---|
| Rhi | Recovered with high relapse risk, following treatment non-completion or self-cure |
| R | Long-term, ‘stabilized’ relapse risk |
| Dx | Presented for diagnosis |
| Tx | Initiated first line treatment |
| Tx2 | Initiated second line treatment |
| s.infectious | All the compartments contributing to spreading infection |
| s.infectious_wosc | Compartments subject to TB mortality |
| s.prevalent | All compartments constituting prevalent TB |

### Outputs used in model calibration

*Table 2: Auxiliary measures*

| | |
|---|---|
| inc | Incidence rate (all TB, hiv +ve TB and RR-TB) |
| noti | Public sector notification (TB that is HIV-negative, HIV-positive, and on ART) |
| noti2 | RR-TB notification (initiating second line treatment) |
| mort | TB mortality (HIV negative and HIV coinfected mortality) |

### Model functions

*Table 3: Model functions*

| | |
|---|---|
| Model_setup | To define all variables, parameters and assign their default values, as well as specifying posterior densities corresponding to input data |
| get_address | To construct lookup tables for compartment numbers relating to each state variable |
| get_distribution_fns | Function to find log-density functions matching given data (e.g. incidence) and uncertainty intervals |
| make_model2 | Specify the full model in matrix form, given all model parameters |

| | |
|---|---|
| goveqs_basis2 | Calculate local gradient for given values of state variables (used in ODE solver) |
| goveqs_scaleup | To capture linear scaleup of parameters between two time points (e.g. used for linear scale-up of interventions) |
| alloc_parameters | Given a parameter vector x, to allocate parameter values (p: proportion, r: rate) |
| get_objective | Given a parameter vector x, to simulate the model and calculate the log-posterior density |
| Get_calibrations2 | To calibrate the model using MCMC |
| MCMC_adaptive | Adaptive MCMC, using Haario et al |
| goveqs_basis_disruption | As for goveqs_basis, but used during periods of disruption, allowing rates of diagnosis to vary |
| goveqs_scaleup_disruption | As for goveqs_scaleup, but used during periods of disruption, allowing rates of diagnosis to vary |
| Show_model_fits1 | To show calibration results |
| Simulate_forward | Forward projection with different intervention scenarios |
| Figure_final | To plot incidence and mortality projection |
| jbfill | Function to show shaded areas for uncertainty intervals |
| linspecer | To specify color series for plotting |

### Natural history parameters

*Table 4: Natural history parameters, assigned values and references.*

| Symbol | Definition | Assigned values | References/Note |
|---|---|---|---|
| r.progression | Per-capita annual rate of progression from 'fast' latent infection (differentiated by HIV-status, h0, h1, hart) | [0.0826 0.8260 0.1652] | Calibration: Menzies (2018) [1] for hiv-ve and 10 times higher for hiv+ve and with ART its rate reduces by 80% |

| | | | |
|---|---|---|---|
| r.LTBI_stabil | Per-capita annual rate of stabilization from 'fast' to 'slow' latent status (differentiated by HIV-status, h0, h1, hart) | [0.8720 0 0.8720] | Menzies (2018) [1] for HIV-ve and for HIV+ve with ART |
| r.reactivation | Per-capita annual rate of reactivation from 'slow' latent infection (differentiated by HIV-status, h0, h1, hart) | [0.0006 0.0600 0.0120] | Calibration: Menzies (2018) [1] for hiv-ve and 100 times higher for hiv+ve and with ART its rate reduces by 80% |
| r.relapse(1) (ro_lo) | Per-capita annual rate of relapse in first two years after treatment completion | 0.032 | Thomas A et al (2005) [2], Romanowski (2019)[3], Menzies (2009) [4] and Weis (1994) [5], with uniform prior using intervals of $\pm$ 5% |
| r.relapse(2) (ro_hi) | Per-capita annual rate of relapse in first two years after self-cure or incomplete treatment | 0.14 | |
| r.relapse(3) | Per-capita annual rate of relapse >two years after last TB episode | 0.0015 | Most relapse occurs in first two years after recovery: Guerra-Assuncao (2015) [6] |
| r.mort | Per-capita annual rate of background mortality | | Corresponds to average lifespan of 70 years (World Bank 2021) [7] |
| p.imm | Immune protection from reinfection | [0.8 0 0.8] | Assumption, with uniform prior using intervals of $\pm$ 25% |

| | | | |
|---|---|---|---|
| r.Dx | Per-capita rate of offering diagnosis | 52 | Assumption: corresponds to an average of 1 week to arrive at a diagnosis |
| p_MDRrec2015 | Of diagnosed TB with rifampcin resistance, proportion that is recognized as such in 2015 (through DST) | [0.001 0] | It is assumed to be a very low value if data is not available for a country |
| p. Tx_init2 | Proportion of second-line treatment initiation after diagnosis as RR | [0.88 0] | Assumption in absence of country specific data |
| p.SL_trans | Amongst RR-TB incorrectly initiated on FL treatment, proportion that is subsequently transferred to second-line treatment | [0.88 0] | Assumption in absence of country specific data |
| p.Tx_init | Of diagnosed patients, proportion initiating first-line treatment | [1 1] | |
| r.Tx | Per-capita annual rate of first-line treatment completion | 2 | Corresponds to average duration of 6 months |
| r.Tx2 | Per-capita annual rate of second-line treatment completion | 0.5 | Corresponds to average duration of 2 years |
| p.cure | Proportion cure after successful completion of FL treatment | [1 1] | |

| | | | |
|---|---|---|---|
| p.tsrsl | Proportion treatment completion of SL treatment | [0.48 1e-6] | Country specific |
| r.default2 | Per-capita annual rate of treatment interruption during SL treatment (r.default2) in public and private sector | | Calculated using $r.default2 = r.Tx2 * p.tsrsl/(1 - p.tsrsl)$, for values of $r.Tx2,\ p.tsrsl$ given above |
| p.cure2 | Proportion cure after successful completion of SL treatment | [0.5 0] | Taken from country reports where available |
| prm.ART_start | Year of ART initiation | | Country specific |
| HIV_incd | Data for annual HIV incidence: assume HIV burden scaled up linearly from 1980 to first data point in 1990 and then using HIV incidence data till 2019 | | Country specific |
| prm.rHIV | Per-capita rate of HIV acquisition, adjusted in to give model agreement with HIV_incd | | Calibration: Estimated |
| r.self_cure | Per-capita annual rate of TB self-cure | 0.17 (0.13 – 0.21) | Tiemersma et al., (2011) [8] for central value, with uniform prior using intervals of $\pm$ 15% |

### Parameters that are estimated during fitting

*Table 5: Parameters that are estimated within assigned ranges as part of the fitting process*

| Symbol | Definition | Assigned ranges |
|---|---|---|
| r_beta | Infection rate (number of annual infections per case) of DS TB | [0 - 40] |
| rfbeta_mdr | Infection rate of RR-TB relative to DS TB | [0 - 1] |
| rfbeta_hiv | Relative transmission rate for HIV +ve TB patients, relative to HIV -ve TB | [0 - 1] |
| r_sym | Per-capita annual rate of developing symptoms, amongst subclinical TB | [0.1 - 100] |
| p_pu | Proportion of care-seeking visits that are to the public sector | [0 - 1] |
| r_cs (1997) | Rate-of-presentation to care, first care-seeking visit for symptomatic TB in 1997 | [0.1 - 100] |
| rf_cs 2022 | Rate-of-presentation to care in 2022 relative to 1997 | [1 - 10] |
| r_cs2 | Rate-of-presentation to care, second and subsequent care-seeking visits for symptomatic TB | [1 - 24] |
| r_mort_TB | Per-capita annual rate of TB mortality while untreated differentiated by HIV-status (hiv-ve and hiv+ve) | 1/6*[0 - 2; 0 - 100] |
| p_Dx | Probability of successful TB diagnosis and treatment initiation per care-seeking visit (in public and private sector) | [0.75- 0.9; 0.1- 0.3] |
| p_TX_complete | Proportion treatment completion (used to estimate per-capita annual rate of treatment interruption (r.default) in public and private sector) | [0.75 - 0.95; 0.4 - 0.8] |
| p_MDRrec2022 | Of diagnosed TB with rifampcin resistance, proportion that is recognized as such in 2022 (through DST) | [0 - 1] |
| r_MDR_acqu | Per capita rate of acquired RR/MDR during treatment | [0 - 0.06] |
| r_ART_init | Per-capita rate of ART initiation | [0 - 10] |
| r_HIV_mort | Per-capita annual rate of HIV mortality while untreated | [0 - 10] |
| r_self_cure | Per capita annual rate of self-cure | 1/6*[0.85 - 1.15] |
| p_HIV_relrate | Progression and activation rate among HIV positive, relative to HIV-negative people | [1 - 100] |

### Intervention parameters

*Table 6: Interventions*

| | |
|---|---|
| p.pu | This value increases with the private sector engagement intervention |
| r.cs/ r.cs2 (in the model $\gamma$ / $\tilde{\gamma}$) | Parameter related to case-finding activity |
| p.DX(1), p.DX(2) | To improve diagnosis in public sector and private sector respectively |
| r.progression and r.reactivation | These parameters reduce with preventive measures |
| p.MDR_rec | Increases to increase drug susceptibility testing |
| r.Tx2 | Intervention parameter to reduce the duration of second line treatment |
| p.cure2 | Intervention on second-line treatment success rate |
| r.default(1) | Increase of treatment completion and reduction of ILTFU can be modelled by reducing r.default(1). |
| p.PT_PLHIV | Proportion reduction of progression rate resulting from TPT among PLHIV |
| r.cs3 | Per-capita rate of case-finding amongst sub-clinical TB |
| p.VE(1) | Vaccine efficacy on reduction of susceptibility (pre-exposure protection) |
| p.VE(2) | Vaccine efficacy on reduction of progression to active disease (post-exposure protection) |
| r.vacc | Per-capita annual rate of vaccination |
| r.waning | Per-capita annual rate of waning vaccine immunity |

### Country-specific calibration targets

*Table 7: WHO data used for calibration purposes*

| | |
|---|---|
| popn | Population size in 2022 |
| data.inc_all | Total TB incidence rate in 2000 and 2022 |
| data.inc_h1 | HIV-positive TB incidence in 2022 |
| data.noti | TB notification rate in 2022 |
| data.mort_H0 | HIV-negative TB mortality in 2000 and 2022 |
| data.mort_H1 | HIV-positive TB mortality in 2022 |
| data.sym | Proportion of prevalent TB that has symptoms |
| data.mdr2015 | MDR/RR-TB incidence in 2015 |
| data.mdr2019 | MDR/RR-TB incidence in 2022 |
| data.mdriniTX | MDR/RR-TB cases started on second-line treatment in 2022 |
| data.ART_covg | ART coverage in 2022 |
| data.HIV_prev | Prevalence of HIV in 2022 |
